# From Cause to Recovery: The Influence of Traumatic Brain Injury Mechanisms on Long-Term Functional Independence

**DOI:** 10.64898/2026.07.10.26357252

**Authors:** Matthew J. Beth, Jennifer Marwitz, Nojan Valadi, Niyati Baweja, Harsimran S Baweja

**Author notes:** Corresponding author: Harsimran S. Baweja P.T. Ph.D., Director of the Auburn University Physical Therapy Program, Director of the SensoriMotor and Rehabilitation Technology Neuroscience Lab School of Kinesiology, Auburn University, 301 Wire Road, Auburn, AL, 36849. These authors contributed equally to this work (Matthew J. Beth, Harsimran S Baweja).

## Abstract

**Background/Objectives:** This systematic review examines how different mechanisms of Traumatic Brain Injury (TBI) influence post-injury functional independence and aims to clarify whether recovery patterns vary by injury type. A total of 105 studies (n = 59,621) involving adults with TBI were synthesized. These findings can guide clinicians and researchers in predicting outcomes and effectively customizing rehabilitation plans.

**Methods:** A review following PRISMA standards analyzed English-language studies published from 1975 to 2025, assessed functional outcomes using the Functional Independence Measure (FIM) or the Glasgow Outcome Scale-Extended (GOSE), converted them to z-scores, and aggregated them via a random-effects model with inverse-variance weighting to demonstrate their relevance.

**Results:** Recreational TBIs show the highest functional independence (*z* = +1.77), followed by MVAs (*z* = +1.56), with falls (*z* = +0.70) and assault-related TBIs (*z* = −0.12) showing moderate outcomes, and TBIs with penetrating trauma (*z* = −1.15) indicating the most adverse results.

**Conclusions:** TBI mechanisms appear to meaningfully influence long-term post-injury functional independence. Highlighting this can inspire clinicians and researchers to trust these insights to improve prognosis and rehabilitation strategies, underscoring their crucial role in advancing patient care.

## INTRODUCTION

Traumatic Brain Injury (TBI) represents a major challenge in public health worldwide, underscoring its critical importance for medical professionals and researchers alike. Defined as a neural injury caused by an external mechanical force acting on the head [1]. TBI encompasses a remarkably heterogeneous range of injury severities, from a common mild concussion that resolves within days to a catastrophic open-head injury that permanently alters every aspect of an individual’s life. Across all TBI severity levels, it is estimated that 64–74 million TBIs occur globally each year [2], and in 2021 alone, TBIs were responsible for approximately 5.48 million years lived with disabilities [3]. These injuries are often chronic, lifelong burdens, rather than mere acute medical events; often entailing persistent motor, cognitive, and emotional impairments that influence a survivor’s ability to live independently, work, and participate in community life long after discharge from the hospital.

In addition to the lifestyle burdens associated with TBIs, the economic burden is also profound. Individuals with moderate-to-severe TBIs (msTBIs) often necessitate extended hospitalization and intensive inpatient rehabilitation [4], and the total annual cost of TBIs to worldwide healthcare systems exceeds 400 billion USD [1]. Individually, in-hospital costs of a severe TBI can range from approximately 2,100 to ≥ 400,000 USD [5], with the expenses further exacerbated following hospital and/or rehabilitation discharge, as survivors of TBI must manage outpatient rehabilitation, acquiring new adaptive equipment, salary reductions, and the long-term consequences of their newly acquired disabilities. Recognizing the importance of predictive research, identifying reliable indicators of recovery trajectories is crucial for improving clinical decision-making and resource allocation. Such research will support the development of personalized rehabilitation strategies, ultimately reducing the long-term societal and economic strife associated with TBIs.

### Mechanism of Injury

Among the variables influencing recovery trajectories, injury severity has been a central focus. A meta-analysis of 21,050 hospitalized individuals with msTBIs found a statistically significant moderate association between TBI severity and functional outcomes one year post-injury (average effect size *r* = .26), but the analysis also showed marked between-study heterogeneity (*I*² = 93.1%), indicating that severity alone does not fully explain differences in recovery outcomes [6] This limitation is reinforced by longitudinal follow-up research: 24 years post-TBI, fewer than 30% of a sample of survivors of TBI were employed full-time [7], and another study found that at 8 years post-injury, nearly half of their sample reported still experiencing persistent balance difficulties and nearly one-third reported significant ongoing motor deficits [8]. These findings cannot be explained solely by acute severity.

The variability has led researchers to investigate the injury mechanism as a potential independent prognostic variable. TBI mechanisms are extremely diverse. Falls are the most common mechanism of TBI in high-income countries, accounting for nearly half of TBI-related hospitalizations in the United States [9] and 56% of all TBIs in the European CENTER-TBI registry, where fall-related TBIs were associated with notably older populations (median age 74 years) [1]. TBIs from motor vehicle accidents (MVAs) commonly occur among younger adults and are a leading TBI mechanism in low- and middle-income countries [1,10]. Assault-related TBIs are particularly common among incarcerated, adolescent, and veteran populations [11]. Conversely, blast injuries are predominantly observed among military populations. These injuries, involving shockwave-related pressure changes, can produce complex, diffuse neural injuries [12]. TBIs caused by recreational and sports-related activities affect younger, more physically active populations and are frequently reported to improve awareness and diagnostic practice [13].

Because TBI mechanisms tend to cluster across demographic groups and yield characteristic injury profiles, comparing functional outcomes across TBI mechanisms is inherently difficult. For example, falls are disproportionately common among older adults who often have a lower neuroplastic potential during their recovery [14], greater comorbidity, higher pre-injury frailty, and more reserved acute healthcare compared to younger individuals with similar TBI severity [1]. Conversely, MVAs often occur in younger individuals whose brains possess a greater capacity for neural reorganization and recovery [14,15]. Likewise, TBIs from recreational activities often occur in younger and more physically active populations. This association between TBI mechanism and normative age, which researchers have labeled as “age-mechanism collinearity”, significantly complicates interpretation, because any unadjusted comparison between TBI mechanisms will reflect, to a certain degree, the normative age of each cohort [16].

### Current Evidence and Its Divergence

Critical knowledge in this area stemmed from the IMPACT (International Mission for Prognosis and Analysis of Clinical Trials in TBI) study, which examined data from more than 8,700 individuals and found that traffic-related TBIs and those from recreational activities were associated with significantly better odds of more favorable long-term outcomes compared to falls or assaults [17]. Notably, the IMPACT results revealed a paradox that has recurred over time in the TBI literature: despite causing more severe acute injuries, MVAs are associated with better long-term recovery outcomes than those from falls, which generally present with less severe acute injuries. A later epidemiological study across five Central European countries reinforced this pattern, showing that 52% of individuals with traffic-related TBIs achieved favorable long-term outcomes, compared with only 42% of those injured by falls [18], thereby strengthening the argument that mechanisms and severity can behave in opposite ways.

The paradox has sparked great debate across the field. One perspective holds that differences in cohort demographics can explain the apparent injury mechanism effect, as falls tend to have worse outcomes simply because older adults fall and experience poorer recoveries, regardless of injury type [16]. The stance was supported by several other studies showing that age is consistently one of the strongest independent predictors of post-TBI functional outcome [19]. An opposing viewpoint is that the TBI mechanism itself exerts biological effects beyond those of normative aging (e.g., focal contusions from fall-related TBI in older adults demonstrate an injury phenotype distinct from that of younger individuals with diffuse axonal injuries from high-speed MVAs, with distinct neuroplastic implications [20]). Evidence reinforcing the notion is found in studies involving working-age adults, which, statistically controlling for age, still observed mechanism-specific outcome differences [17,18,21], as well as in findings that mechanism-related differences tend to become more pronounced across the lifespan, rather than being evident only at rehabilitation discharge [21]. This pattern is difficult to explain solely in terms of age-related biology.

Violence-related and penetrating TBIs add a layer of complexity to this discussion, because their poorer prognosis appears to be mediated at least partly by psychosocial rather than purely biological factors. Studies have shown that survivors of violence-related TBIs have significantly poorer employment and community reintegration one year post-injury than those injured in falls or MVAs, even though mechanism-related differences were not evident at rehabilitation discharge [Bushnik], suggesting that the social consequences of acts of violence, such as Post Traumatic Stress Disorder (PTSD), substance abuse, financial difficulties, and poor social support, may exacerbate the biological effects of TBIs over time. A UK cohort study demonstrated that assault survivors reported substantially worse quality of life and higher rates of PTSD than fall survivors [22]. However, another study did not find any functional outcome differences between penetrating and closed head violence-related TBIs among survivors [23], suggesting the possibility that the disadvantages associated with violence-related TBIs may be mediated by the psychosocial context of the violent act, rather than from the penetrating nature of the injury itself. Latent class and latent profile analyses provide additional detail: open-head injuries and delayed rehabilitation admission predicted worse motor recovery trajectories [24]. Meanwhile, a different study identified a distinct subgroup of the most severely impaired survivors of TBI, composed of individuals with open-head injuries and profound consciousness impairments. The subgroup disproportionately sustained violence-related injuries and demonstrated the poorest long-term FIM and GOSE scores [25].

Conversely, the literature shows that recreational TBIs are associated with greater functional independence [17], which may be explained by a combination of affected individuals tending to a younger age with higher baseline physical fitness, who predominantly sustain diffuse injuries, and often have structured post-TBI care environments. Individuals experience a critical period of elevated neuroplasticity during the first six months after TBI onset. The time is characterized by increased axonal sprouting, dendritic remodeling, synaptogenesis, upregulation of N-methyl-D-aspartate (NMDA) receptors, and the downregulation of Gamma-Aminobutyric Acid type A (GABAA) receptors [26,27]. The critical period is particularly well utilized by younger populations, who have greater biological reserve, which may explain the sustained recovery advantage observed in recreational TBI groups over extended follow-ups [14,15].

### Gaps in the Literature

Despite an increasing body of evidence, multiple key gaps in the literature remain. Too few studies have explicitly examined mechanism-age interactions, leaving the unanswered question of whether the effects of TBI mechanisms extend beyond age. Literature is also significantly heterogeneous in the instrumentation used to collect functional outcome data: some studies use the Functional Independence Measure (FIM) [28,29], while others use the Glasgow Outcome Scale–Extended (GOSE) [30,31], and still others use different domain-specific measures of cognition, employment, or community reintegration, complicating synthesis across studies without standardization. No previous systematic review has combined outcomes across common TBI mechanisms within a single analytic framework using a common metric, leaving clinicians without a clear evidence-based comparison of mechanism-specific functional prognosis. Furthermore, populations from low- and middle-income countries are underrepresented [1,10], despite MVAs and violence accounting for a disproportionately large portion of the TBI burden in the socioeconomic settings, and evidence for mechanism-specific rehabilitation interventions is limited [32].

### Objective and Hypotheses

This systematic review addresses these gaps in the literature by synthesizing evidence from 105 studies (*n* = 59,621 participants) on the relationship between TBI mechanisms (e.g., falls, MVAs, assaults, penetrating injuries, and recreational injuries) and post-TBI functional independence in adults, measured by either the FIM or GOSE. To facilitate direct comparisons across instrumentation differences, outcomes were standardized to z-scores (z_FIM_ and z_GOSE_), enabling clinicians and researchers to interpret results within a common metric. The review followed the Preferred Reporting Items for Systematic Reviews and Meta-Analyses (PRISMA) guidelines, and pooled z-score estimates were calculated using a random-effects model with inverse-variance weighting.

Based on the theoretical and empirical literature summarized above, three hypotheses were tested to inform mechanism-specific rehabilitation strategies. Consistent with previous literature, we expected that individuals with TBIs from falls, assaults, and penetrating injuries would show poorer functional independence compared to individuals injured from MVAs or recreational activities [17,18,21,22]. Second, we expected to observe an association between penetrating injuries and the poorest outcomes across all mechanism groups, given the relation of these injuries to complex focal neural damage, delayed rehabilitation, and the psychosocial burden associated with acts of violence [24,33]. Third, we expected recreational TBIs to be associated with the highest functional independence scores, reflecting the physically active demographic and biological profile of this population and the common diffuse, mild-to-moderate nature of sport-related TBIs [14,17]. These hypotheses aim to guide targeted rehabilitation interventions based on mechanism-specific outcomes, thereby advancing personalized care approaches.

## MATERIALS & METHODS

### Protocol

This systematic review followed the Preferred Reporting Items for Systematic Reviews and Meta-Analyses (PRISMA) guidelines, demonstrating a comprehensive approach that aims to instill confidence in researchers and clinicians about the rigor of our methodology.

### Eligibility Criteria

#### Population

The researchers included studies of adults aged 18 years or older, regardless of Traumatic Brain Injury (TBI) severity classification (mild, moderate, or severe). The researchers excluded studies involving pediatric populations or non-traumatic brain injuries (e.g., acquired brain injuries and anoxic injuries). Studies examining individuals with mechanisms of TBI from falls, motor vehicle accidents (MVAs), assaults, blast injuries, gun violence, and sports-related trauma were eligible for inclusion in this study.

#### Outcome Measures

Researchers and clinicians evaluate functional independence in numerous contexts. Functional independence is described as one’s ability to function safely and independently within one’s own context. Independence is determined by how well a person performs activities of daily living (ADLs) autonomously. Tools for measuring functional independence include standardized measures such as the Functional Independence Measure (FIM) and the Glasgow Outcome Scale-Extended (GOSE). Therefore, the scores of both tools were the primary focus of this review. Secondary outcomes included cognitive tests, such as the Trail-Making Test, and measures of return to work, return to play, and community reintegration.

The FIM comprises 18 items, divided into motor (13 items) and cognitive (5 items) subscales, scored on a 7-point ordinal scale (total FIM scores range from 18 to 126) that quantifies one’s level of independence [34]. Higher scores indicate greater functional independence. It is a validated and widely used tool in TBI rehabilitation research. GOSE is an 8-point ordinal scale used to classify global outcomes after TBI [31,35]. It expands upon the original 5-level Glasgow Outcome Scale by subdividing several categories into “lower” and “upper” bands, yielding scores ranging from 1 (death) to 8 (upper good recovery) [35]. Similar to FIM, higher scores indicate greater functional independence: death (1), vegetative state (2), lower or upper severe disability (3–4), lower or upper moderate disability (5–6), and lower or upper good recovery (7–8) [35]. Researchers collect GOSE data through semi-structured interviews that assess changes in functioning relative to pre-injury levels across domains such as consciousness, self-care, mobility, work or school, and social participation. [35]. It is one of the most widely used functional outcome measures in TBI research [31,36]. Both the FIM and the GOSE are susceptible to experiencing ceiling effects. To address this, median differences were used to analyze the data. Additionally, several studies used odds ratios to dichotomize results into higher- and lower-score categories.

### Study Design

This systematic review primarily focused on observational studies (including cohort and cross-sectional designs) and a limited number of randomized controlled trials (RCTs) that investigated the influence of TBI recovery on functional outcomes.

#### Language and Timeframe

This review included studies published in English between March 1975 and February 2025, accounting for original measurement documentation while also ensuring current clinical relevance. Additionally, the extended time span was intended to adequately capture longitudinal changes.

#### Sources and Search Strategy

A thorough literature search identified 149 records from MEDLINE (n = 19), CINAHL (n = 25), PsycINFO (n = 39), ScienceDirect (n = 25), JSTOR Journals (n = 4), APA PsycArticles (n = 6), Science Citation Index Expanded (n = 25), and Academic Search Premier (n = 6) electronic databases were identified using search terms related to (1) Traumatic Brain Injury, (2) etiology/mechanism of injury, and (3) long-term functional independence. Using Boolean operators (“And”, “Or”), the search terms for the Traumatic Brain Injury component of the review included combinations of: “traumatic brain injury”, “TBI”, “head injury”, “neurotrauma”, “open head injury”, and “closed head injury”. For etiology/mechanism of injury, the following search terms were used in combination with “Traumatic Brain Injury“: “fall”, “motor vehicle accident”, “road traffic accident”, “car crash”, “blast”, “assault”, “violence”, “gunshot wounds”, “sports-related”, “penetrating injury”, “industrial accident”, “work-related”, “recreational”, as well as descriptors like “mechanism of injury”, “cause”, and “etiology”. Lastly, for the functional independence facet of this review, search terms included “functional independence”, “independent living”, “community living”, “activities of daily living”, “ADL”, “instrumental activities of daily living”, “IADL”, “participation”, “social participation”, “community involvement”, “involvement”, “community reintegration”, “employment”, “return to work”, “return to school”, “long-term”, “longitudinal”, “follow-up”, “outcome”, and “recovery”. “Long-term” was operationally defined in the search strategy by using terms that indicated an extended time horizon (e.g., “long-term”, “follow-up”, “longitudinal”, “after injury”, “months”, “years”) and by applying study inclusion criteria during screening rather than by setting a single cutoff within the query itself. In each database, these three blocks were combined as follows: (TBI terms) AND (etiology/mechanism terms OR etiology synonyms) AND (functional independence terms OR independence/outcome synonyms). Filters for human participants and study type were applied where available within each database, and additional filters (e.g., article publication dates) were applied only at the screening stage. After screening the 149 records and determining eligibility, the systematic review included 105 records in this analysis.

#### Selection Process

Records identified through database searches were uploaded to Covidence systematic review software (Veritas Health Innovation, Melbourne, Australia) for screening and management. This systematic review considers studies that report outcomes using the FIM or GOSE, as well as articles on the instruments’ psychometric properties. To capture a comprehensive, holistic depiction of TBI mechanism-specific recovery patterns, the review also considers articles on post-injury neuroplasticity, biomarkers, and well-being profiles. Duplicate records were identified and removed before screening. Record titles and abstracts were screened for eligibility by the first author in accordance with the inclusion and exclusion criteria. Studies were excluded if they involved non-TBI populations, pediatric populations, or non-human populations, or if they did not report functional outcomes using either the FIM or GOSE.

Full-text articles were retrieved for records meeting the screening criteria. The same eligibility criteria were applied to full-text screening, including adult TBI populations (18+ years old), the specification of injury mechanism and/or injury severity, and the reporting of functional outcomes using the FIM or the GOSE. Articles were excluded at the full-text stage if they did not report relevant functional outcome measures, involved pediatric, non-human, or non-TBI populations, or did not have copies in English available. The rationale for exclusions at the full-text stage was documented in Covidence.

The database search initially yielded 149 records, and the researchers uploaded all retrieved records to Covidence. After removing 32 duplicate records, 117 studies underwent title and abstract screening. Following title and abstract screening, 12 records were excluded for not meeting the eligibility criteria, leaving 105 articles for full-text review and inclusion in the final synthesis (see Table S1). The selection process is presented in a PRISMA flow diagram (see Figure S1).

#### Data Extraction

The researchers extracted data on study design and characteristics (author, year, and country), sample size, participant demographics (age, gender, residence location, education level, and occupation type), intervention protocols and instrumentation, TBI mechanism and severity, inclusion and exclusion criteria, outcome measures assessing functional independence (including FIM and GOSE), and follow-up duration. In addition to Covidence, the researchers extracted data using Microsoft Excel (Microsoft Corporation, Redmond, WA, USA; Version 16.105.2).

#### Instrumentation Differences

Studies using the same outcome measures could be easily compared. However, the reviewed studies demonstrated considerable heterogeneity in the instrumentation used to evaluate functional outcomes. This review focuses on studies that use either the FIM or the GOSE. For comparison of the measures, data obtained from different instruments were standardized by calculating z_FIM_ and z_GOSE_:

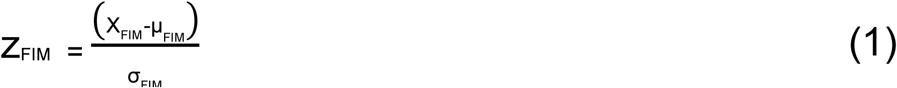

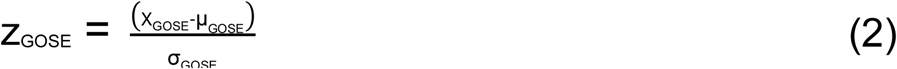

where X denotes the individual score on each respective instrument, μ represents the group mean score on each instrument, and σ denotes the standard deviation of the scores on each.

#### Quality Assessment

The researchers evaluated the studies for quality and assessed them for risk of bias. They evaluated the quality of evidence for each study by examining the levels of evidence outlined in Sackett’s framework [37]:

- Level I: Meta-analyses, systematic reviews, and RCTs.
- Level II: Nonrandomized, two-group cohort studies.
- Level III: Nonrandomized, one group (pre/post-test).
- Level IV: Single-subject and case series studies.
- Level V: Case reports and narrative literature reviews.

Risk of bias (RoB) was evaluated using validated tools specific to each study design. Observational studies, including cohort and cross-sectional study designs, were assessed with the Newcastle–Ottawa Scale (NOS) [38]. The tool examines methodological rigor across three domains: study population selection, between-group comparability, and outcome assessment (for cohort studies) or outcome collection (for cross-sectional studies). Scores were assigned for each domain according to predefined criteria, with higher total scores indicating a lower RoB. Table S2 presents the NOS’s criteria for assessing RoB in observational studies.

To examine the effects of various therapeutic practices on functional recovery among individuals with different TBI mechanisms and distinct recovery patterns, this systematic review included 2 RCTs. The couple of included RCTs were assessed using the Cochrane Risk of Bias Tool, version 2.0 (RoB 2) [39], which evaluates potential bias across five domains: the randomization process, deviations from interventions, missing data, outcome measurement quality, and selective reporting of results. Domains were rated as “low RoB,” “some concerns,” or “high RoB,” and an overall risk-of-bias assessment was made in accordance with the Cochrane recommendations. Evaluation of the two RCTs is presented in Table S3.

All RoB evaluations were performed by the lead author (MJB) using standardized assessment procedures and published guidance for each instrument to enhance consistency and reproducibility. RoB ratings were used to contextualize the interpretation of each study’s results, but did not exclude studies from the review.

#### RoB from missing results (Reporting Bias)

Bias due to missing results was evaluated at the synthesis level by the lead author (MJB). In RCTs, the relevant domain of the RoB 2, addressing bias arising from missing outcome data and selective reporting, was applied. Judgments were informed by comparisons between outcomes specified in study methods or trial registrations.

Observational studies were assessed with the NOS. Reporting bias was evaluated qualitatively by reviewing the completeness and consistency of outcome reporting across studies. This included assessing whether functional outcome measures were reported for applicable study participant groups. Across study designs, inconsistencies between outcomes outlined in the methods, unexplained omissions, and incomplete reporting of outcome data were systematically documented.

#### Exploration of Heterogeneity

Given the significant heterogeneity across the analyzed studies in study design, participant characteristics, injury severity, functional independence outcome measures, and follow-up duration, this heterogeneity was explored qualitatively and through subgroup stratification by key study characteristics.

To examine whether functional independence outcomes differed across injury mechanisms, studies were grouped by injury mechanism (e.g., falls, motor vehicle accidents, assaults, penetration injuries, and recreational activities). To facilitate comparisons across studies, functional outcome measures from the FIM and the GOSE were transformed using z-scores.

Mechanism-specific pooled z-score estimates were generated using a random-effects model based on the DerSimonian-Laird method, with inverse-variance weighting across studies contributing data to each mechanism category. Between-study heterogeneity was quantified using the *I²* statistic and is reported alongside the pooled estimate and 95% confidence interval for each mechanism row in the results synthesis. When the same study contributed data to more than one mechanism category, those subgroup-level estimates were treated as independent contributions, an assumption that is acknowledged as a methodological limitation in the Discussion.

Other sources of heterogeneity were examined descriptively by juxtaposing results across subgroups characterized by TBI severity, age, and follow-up duration. Patterns of heterogeneity were interpreted based on the consistency of findings within mechanism-specific groups and across levels of injury severity.

## RESULTS

### Study Selection

The initial database search identified 149 records. After removing duplicates (*n* = 32), the researchers screened 117 studies based on their titles and abstracts. The researchers assessed the records for eligibility and excluded 12 full-text articles that did not meet the inclusion criteria. Following the full-text review, the researchers determined 105 studies (see Table S1) met the inclusion criteria and were included in the final synthesis. The Preferred Reporting Items for Systematic Reviews and Meta-Analyses (PRISMA) flow diagram (see Figure S1) illustrates the study selection process, detailing the number of studies screened and the final number included.

### Study Characteristics

Table S4 presents an example of the study characteristics data extraction conducted for seven of the 105 included studies. Studies differed by design (e.g., prospective cohorts, meta-analyses, epidemiological studies), geographical location, sample size, Traumatic Brain Injury (TBI) mechanisms and/or severity measures, and follow-up data duration. Included studies evaluated adult populations with TBI and post-TBI functional outcomes, particularly those related to TBI mechanisms or severity.

### Functional Findings by TBI Mechanism

In line with the analytical approach, Functional Independence Measure (FIM) and Glasgow Outcome Scale-Extended (GOSE) scores were standardized to z-scores (z_FIM_ and z_GOSE_) to allow comparison across studies using different methodologies and equipment.

For analytical purposes, TBI mechanisms were grouped as either falls, motor vehicle accidents (MVAs), penetrating injuries (gunshot wounds, explosions, etc.), assaults, or recreational activities. In line with the analytical approach, FIM and GOSE scores were standardized as z-scores (z_FIM_ and z_GOSE_) to enable comparisons across studies using different methodologies and equipment.

Across the 105 included studies, approximately *n* = 59,621 participants contributed functional outcome data. Of these, *k* = 46 studies reported FIM measures exclusively, *k* = 52 reported GOSE measures exclusively, and *k* = 7 reported measures from both instruments. Z-scores calculated from the extracted data and pooled Z-scores are displayed in Table S5. Additionally, the pooled z-scores for each TBI mechanism group are depicted in a forest plot in Figure S2.

#### Falls

Number of studies *k* = 82; Sample *n* = 11,309.

Falls are associated with moderate standardized functional outcomes/independence across studies, with z_FIM_ values ranging from +0.87 to +1.28 and z_GOSE_ values ranging from 0.00 to +0.63.

The pooled z-score estimate indicated a slightly lower level of functional independence among individuals with fall-related TBIs than among those with other TBI mechanisms (mean *z* = +0.70, 95% CI: [0.66, 0.74]).

#### Motor Vehicle Accidents (MVAs)

Number of studies *k* = 79; Sample *n* = 8,964.

MVAs are disproportionately more common among younger adults. Outcomes varied widely and were heavily contingent upon injury severity and the presence of polytrauma. Collectively, MVAs demonstrated relatively higher standardized functional outcomes than most other mechanisms, with z_FIM_ values ranging from +1.64 to +2.08 and z_GOSE_ values ranging from +0.63 to +1.88.

Pooled estimates indicate greater functional independence (mean *z* = +1.56, 95% CI: [1.52, 1.60]), despite the cohorts often exhibiting higher initial injury severity levels.

#### Penetration Injuries (gunshots, blast injuries, etc.)

Number of studies *k* = 92; Sample *n* = 10,443

Penetration injuries account for a large portion of TBIs caused by acts of violence and are common among military populations. The penetration injuries group demonstrated the poorest functional outcomes across all TBI mechanism groups. Their z_FIM_ values ranged from −2.07 to +0.23, and their z_GOSE_ values ranged from −1.88 to +0.42.

The pooled estimate confirmed significantly poorer outcomes (mean *z* = −1.15, 95% CI: [−1.21, −1.09]), reflecting the most suboptimal functional outcomes across all TBI mechanism groups.

#### Assaults

Number of studies *k* = 88; Sample *n* = 9,990.

TBI severity strongly influenced the functional outcomes of the assault group, which varied across studies. Therefore, assault-related TBIs demonstrated heterogeneous outcomes, with z_FIM_ values ranging from −1.11 to +0.62 and z_GOSE_ values ranging from −0.63 to +0.63.

The pooled estimates (mean *z* = −0.12, 95% CI: [−0.18, −0.06]) indicate functional outcomes that are relatively comparable to those of the falls group but with greater variability.

#### Recreational Activities

Number of studies *k* = 81; Sample *n* = 9,915.

The researchers grouped TBIs sustained during athletic events and leisure activities together as the “recreational injuries” group. The recreational injuries group demonstrated the greatest functional outcomes. The groups’ z_FIM_ values ranged from +1.81 to +2.13, and their z_GOSE_ values ranged from +1.25 to +1.88.

The pooled estimate indicated superior functional outcomes for the group across all TBI mechanism groups (mean *z* = +1.77, 95% CI: [1.74, 1.80]).

## DISCUSSION

To inform post-injury prognostication and clarify how the etiology of Traumatic Brain Injury (TBI) differentially influences functional recovery, we assessed the impact of TBI mechanisms on post-injury functional outcomes and independence. We conducted a comprehensive literature review across multiple electronic databases, identifying 149 records, of which 105 remained after screening and eligibility review. We extracted data from the included studies on study characteristics, participant demographics, TBI mechanism, injury severity, and functional independence measures from either the Functional Independence Measure (FIM) or the Glasgow Outcome Scale-Extended (GOSE). Our review found that falls, which are commonly observed among older adults, were often associated with poorer functional outcomes. Conversely, Motor Vehicle Accidents (MVAs) were often associated with superior motor functional recovery.

Penetration injuries, often caused by acts of violence and gun violence, were associated with poorer motor function and greater psychological difficulties. Individuals with TBIs sustained during recreational activities and athletic participation demonstrated the greatest functional independence among the mechanisms assessed. Collectively, our findings support the study hypothesis that individuals with TBIs caused by falls, assaults, and penetration injuries, often resulting from gunfire and other acts of violence, would demonstrate poorer functional independence than individuals with TBIs caused by other mechanisms.

Global estimates indicate that falls are the most common mechanism of TBI [1]. In high-income countries (HIC), falls are the predominant mechanism of TBI, particularly among older populations. Conversely, low- and middle-income countries (LMIC) exhibit relatively more MVAs and violence-related injuries [1]. For example, the European CENTER-TBI registry (HIC) found 56% of TBIs were due to falls, 71% of which were ground-level; these fall-related cases involved much older patients (median age 74 years vs. 42 years) and a higher proportion of women than high-energy injuries [1]. Conversely, in Uganda (LMIC), 88.9% of unintentional TBIs were from MVAs, only 11.1% were from falls, and 97% of intentional TBIs were assaults [10].

Demographic differences cause distinct functional outcomes. Falls are common among older adults, and older age is independently associated with poorer functional recovery after TBI [1,19]. In this review, fall-related injuries were associated with poorer functional independence, primarily because older populations are commonly affected by falls. In contrast, penetration injuries (often resulting from violence-related injuries) frequently affect younger adults (predominantly males and often involve alcohol consumption or other substances [1]). Functional independence post-TBI greatly varies based on the injury severity of survivors of assault and violence-related TBI, with severe injuries having worse long-term functional recovery and functional outcomes than those injured by falls. Exemplary of this, in a cohort in the UK, assault victims reported notably worse quality of life and Post Traumatic Stress Disorder (PTSD) symptoms in comparison to survivors of falls [22]. Similarly, an extensive U.S. TBI Model Systems study observed that after one year post-injury, violence-related TBI survivors experienced higher rates of unemployment and poorer community integration compared to those injured from MVAs and falls; the success of individuals with fall-related injuries landed between the most successful (MVAs) and the least successful (violence) groups [21]. Notably, the study did not observe differences in TBI mechanism at rehabilitation discharge, suggesting that TBI mechanism has a greater influence on longer-term recovery than on acute FIM measures [21]. (In another analysis, even the subgroup of penetrating versus non-penetrating violence-related injuries showed no FIM differences among survivors [40], suggesting other factors may mitigate the effect of TBI mechanisms). Ultimately, violence-related TBIs are associated with poorer functional independence than unintentional injuries like falls, even after accounting for injury severity [21,22]. However, studies report mixed results and often produce inconsistent findings.

### Disentangling Mechanism from Age

A central interpretive caveat of this review is that the mechanism of injury and age are strongly collinear in the TBI population. Falls cluster disproportionately among older adults, MVAs and recreational injuries are more common among younger adults, and penetration and assault-related injuries often affect a predominantly young-to-middle-aged male demographic [1,11,28]. Because age is independently associated with poorer functional recovery after TBI, any unadjusted comparison of outcomes across mechanisms will inevitably reflect, at least in part, the age structure of each mechanism cohort. Age-related factors such as reduced neuroplastic capacity [14], higher comorbidity burden, pre-injury frailty, baseline dependence [28], and more conservative acute management [1] may all contribute to poorer recovery. Therefore, the poorer fall-related outcomes observed in this review cannot be attributed solely to the biomechanics of low-height falls. They may also reflect that many individuals who sustain fall-related TBIs begin recovery with lower baseline function and a narrower biological reserve.

Several lines of evidence within the included literature, however, suggest that the mechanism may exert an effect beyond age alone. First, studies that restricted analyses to working-age adults or statistically adjusted for age continued to identify mechanism-specific differences, with violence-related and fall-from-height injuries predicting poorer one-year outcomes even after adjustment [30,37,38]. Second, mechanism-specific differences appear to emerge more clearly after rehabilitation discharge than during the acute rehabilitation period, a pattern that is difficult to explain by age alone. If age were the only driver, age-related biological constraints would be expected to influence outcomes across the recovery window rather than becoming more apparent over time. Third, the relationship between initial injury severity and long-term outcome appears to differ by mechanism. Falls are often associated with less severe acute presentations but poorer long-term outcomes, whereas MVAs may present with greater initial injury severity yet demonstrate more favorable recovery trajectories [38]. This pattern suggests that mechanism-specific injury phenotypes, such as focal injury and comorbidity-linked vulnerability in fall-related TBI versus diffuse injury patterns with greater recovery potential in some high-energy mechanisms, may contribute to long-term functional outcomes.

We therefore interpret the mechanism-outcome associations in this review as reflecting a composite of three interacting factors: the biomechanical signature of the injury, the demographic and clinical profile of the individuals who sustain it, and the care pathway they encounter. These findings should not be interpreted as evidence of a pure mechanism effect. Future studies should report outcomes stratified by mechanism within age bands (e.g., 18–45, 46–65, and 66+ years) and, where sample sizes permit, formally test mechanism-by-age interactions. Until such analyses become routine, clinicians should use the mechanism of injury as one component of prognosis rather than as a stand-alone predictor. This is especially important for patients whose age places them outside the typical profile for their mechanism group, such as an older adult injured in a high-speed MVA or a younger adult injured by a ground-level fall.

#### Implications

##### Clinical Implications

The study findings have paramount practical implications. First, understanding the effects of TBI mechanisms can help refine prognoses and discharge planning in rehabilitation settings. Individuals with fall-related TBIs may have an uncertain forecast for recovering independence post-injury due to an increased likelihood of baseline dependency. This study shows that falls result in relatively moderately poor motor functional outcomes, with a pooled mean z-score of +0.70, directly contributing to increased baseline dependency. Therefore, individuals may require extended rehabilitation, implementation of fall-prevention measures, and additional home safety protocols to mitigate the difficulties they are likely to face. Conversely, younger individuals with TBIs from MVAs may be expected to achieve physical recovery more rapidly, as MVAs have a mean *z* = +1.56. However, they may face long-term challenges in community reintegration, mental health, and employment. Previous research has shown that neuroplasticity is significantly elevated in younger individuals than in older individuals [14]. The biological advantages of the youthful brain would increase one’s potential for recovery from physical impairments over time. Meanwhile, previous work has shown that a TBI interrupts key developmental milestones, leading to their psychological underdevelopment and compromising their mental health and community involvement [41,42]. Therefore, rehabilitation plans for violence-related TBIs should incorporate cognitive and behavioral therapies, substance use counseling, and/or other social supports. Using targeted interventions for individuals with violence-related TBIs, tailored to address their disproportionately higher risk for poor community integration and unemployment, is recommended [21].

##### Neuroplasticity and Neurorehabilitation

Understanding how TBI mechanisms affect functional independence impairments and quickly learning prognostic information from the injury site enables clinicians to rapidly design and implement aggressive, tailored therapeutic plans that target areas that may be disproportionately difficult for individuals with a particular TBI mechanism.

Expediting the application of these interventions is paramount for this population, as survivors of neurotrauma experience a time referred to as the “critical period” (or “critical window”) of optimal neuroplasticity and neural healing during the first six months post-injury. A cascade of cellular and molecular processes characterizes the period, including increased axonal sprouting, dendritic remodeling, and synaptogenesis, as well as upregulation of N-methyl-D-aspartate (NMDA) receptors and downregulation of Gamma-Aminobutyric Acid type A (GABAA) receptors [26,27]. The critical period offers survivors of TBI the best opportunity to recover from their disabilities. However, this beneficial time is short-lived, and the critical window gradually closes between 6 and 24 months post-injury. Therefore, it is imperative to administer therapeutic protocols as soon as possible, before this time ends, enabling survivors of TBI the ability to leverage the biological advantages of this time.

The age of the survivor of TBI and the TBI mechanism also significantly affect how individuals experience their critical period. Research has shown that the brains of younger individuals are more plastic, undergoing neural healing and functional improvements over longer periods than those of older individuals [14,15]. Additionally, individuals with diffuse axonal injuries from MVAs will demonstrate a longer recovery trajectory than someone with a focal penetration injury [20]. Therefore, the expedition of implementing mechanism-specific tailored therapies is particularly important for older individuals and/or those with focal penetration injuries.

#### Strengths and Limitations

##### Strengths

The review employed a comprehensive search strategy across multiple electronic databases, including MEDLINE, CINAHL, and PsycINFO, and adhered to the Preferred Reporting Items for Systematic Reviews and Meta-Analyses (PRISMA) standards. Complying with the standards ensures adequate methodological rigor, transparency, and reproducibility. Using a systematic review program (Covidence) streamlines study selection and mitigates possible human error in screening and data management. This review included diverse study designs and drew from studies conducted in multiple countries, supporting some breadth of generalizability across rehabilitation settings; however, the English-only inclusion criterion and the limited representation of low- and middle-income countries (acknowledged in the limitations below) constrain the extent to which these findings can be generalized to all global TBI populations. Additionally, the inclusion of studies published from 1975 through 2025 provides a comprehensive long-term perspective, capturing longitudinal trends in functional outcomes following TBI.

##### Limitations

The limitations of this systematic review include heterogeneity in study designs, outcome measures, and data-collection frequency, which complicates comparisons across studies. Existing literature is limited by the small number of studies that stratify functional outcomes by TBI mechanisms, and those that do may report outcomes only for a select group of mechanisms. Additionally, there is significant heterogeneity in the analytical approaches employed and instrumentation used, which may alter the data collected. Researchers inconsistently adjust for confounding variables, including age and comorbidities, and underrepresent low- and middle-income countries in the literature. Lastly, excluding non-English studies combined with the aforementioned flaws may limit comprehensiveness and generalizability. The limitations highlight the need for standardized methodologies and inclusive research. A further methodological limitation is that title and abstract screening, full-text eligibility decisions, and risk-of-bias assessment were performed by a single reviewer (MJB) without independent duplicate review. While standardized assessment procedures and predefined eligibility criteria were applied throughout, the absence of dual review may have introduced selection or rating bias. Future updates to this review should incorporate independent dual screening and risk-of-bias ratings, with disagreements resolved by consensus or by a third reviewer, in line with current PRISMA and Cochrane guidance.

#### Future Directions

Addressing the limitations identified in the literature requires targeted research efforts. Prospective cohort studies should compare functional trajectories of individuals grouped by TBI mechanism, with a sufficient sample size to adjust for age, severity, and comorbidities. Studies should uniformly use standardized outcome measures (e.g., FIM or GOSE) at multiple follow-up time points to capture functional outcomes at discharge and assess long-term outcomes. Therapeutic intervention trials should explore whether individualized rehabilitation (e.g., adding psychological therapeutic services for violence-related TBIs or fall prevention for older individuals) improves functional independence. Additionally, future research should examine psychosocial outcomes and quality of life alongside the FIM or GOSE, as studies suggest that perceived recovery may vary by TBI mechanism, even when physical function is similar across individuals [22]. Finally, improving socioeconomic and environmental data is necessary; specifically, understanding how return-to-work processes may differ after assaults versus after falls could guide the allocation of vocational rehabilitation resources. Lastly, a list of potential future research avenues and unanswered research questions is below.

##### Recommendations for Future Work

The following are two areas identified for potential growth:

###### (1) Classify Outcomes by Mechanism of Injury

Researchers should design studies that specifically compare functional outcomes (e.g., FIM, GOSE) stratified by TBI mechanism (e.g., falls, assaults, MVAs) to identify mechanism-specific recovery patterns.

###### (2) Incorporate Longer-Term Follow-Ups

Research should examine longitudinal variables by following up with individuals beyond the acute phase, as TBI survivors have demonstrated continued progress for years post-injury. Current studies often employ follow-up durations of 1-8 years. Conducting future studies spanning more than 10 years would enable researchers to assess the longevity and sustainability of functional independence and to identify late-emerging improvements or difficulties.

##### Remaining Questions

Below are three research questions that remain unanswered:

1. What influence does the mechanism of TBI have on short- and long-term functional independence outcomes among individuals from different socioeconomic statuses and healthcare settings?
2. Can identifying the mechanism of TBI inform earlier, individualized rehabilitation strategies, and improve functional independence measures?
3. Are the functional outcomes and independence patterns observed consistent over extended periods (+10 years)?

## Conclusions

Drawing on 105 studies and nearly 60,000 participants over roughly five decades of international research, this systematic review represents the most comprehensive attempt yet to compare functional independence outcomes across TBI mechanisms in a single synthesis. An evident pattern emerged from the evidence: how a person sustains a brain injury appears to significantly influence how well they recover. In descending order, recreational TBIs are associated with the most favorable functional outcomes, followed by motor vehicle accidents (MVAs), falls, assault, and penetrating injuries, which are associated with the least favorable functional recovery. The gradient holds across the literature and becomes increasingly apparent after applying z-score standardization to improve comparability across heterogeneous methodologies. These findings should reassure clinicians and researchers that understanding injury mechanisms can meaningfully inform prognosis and treatment planning.

However, the pattern does not reflect how hard a person hit their head. Acute biomechanical injury severity alone fails to account for the magnitude of differences in functional recovery. Recovery post-TBI unfolds through a myriad of interconnected forces, including the biological makeup of the injury, the individual’s age and health profile, and the social and healthcare environments that support continued recovery after formal treatment ends. Falls exemplify this particularly well. Despite commonly producing injuries with moderate initial severity, fall-related TBIs frequently demonstrate poor long-term functional outcomes, largely attributed to the fact that falls disproportionately affect older adults. They have a diminished capacity for neuroplastic recovery and carry heavier comorbidity burdens. Conversely, TBIs from recreational activities and MVAs often occur in younger populations, who have greater biological reserves for cortical reorganization and adaptation, which helps explain their comparatively favorable recovery trajectories. Penetrating and violence-related TBIs often face challenging and complicated scenarios. They may deal with a combination of their severe focal injury, while also enduring downstream psychosocial consequences, including Post-Traumatic Stress Disorder, substance abuse, economic burdens, and social isolation.

Practically, these findings lay the foundation for treating TBI mechanisms as meaningful inputs for prognostic judgment and rehabilitation planning. Survivors of violence-related and penetrating TBIs often require a more supportive physical and cognitive rehabilitation framework, which may include psychological care, substance abuse support, vocational assistance, and sustained community reintegration services. Across all mechanisms, the evidence reinforces an already-established principle: the early post-TBI critical period, particularly during the first six months post-injury, represents a window of heightened neuroplastic activity when rehabilitation efforts are most likely to yield meaningful gains. Applying these insights can inspire clinicians and researchers to develop targeted interventions that maximize recovery during this vital critical period, fostering hope for improved patient outcomes.

However, confidence in the findings must be tempered by the considerable heterogeneity in the literature. Differences in study design, instrumentation, methodologies, and reporting practices complicate efforts to isolate the independent contribution of the TBI mechanism from the influence of age. Prospective studies that stratify by age, formally test the mechanism-by-age interaction, use standardized outcome measures, and investigate mechanism-specific intervention trials are paramount for advancing understanding and practice. Continued research aligned with these standards can motivate the community to pursue more precise, impactful studies, ultimately leading to more effective and efficient tailored rehabilitation strategies for the millions of people living with TBI worldwide.

## Data Availability

No available data.

## Author Contributions

Conceptualization, M.J.B. and H.S.B.; Methodology, M.J.B.; Data Management, M.J.B.; Analysis, M.J.B.; Writing, M.J.B. and H.S.B; Review and editing, J.M., N.V., N.B., and H.S.B.; Supervision, H.S.B. All authors have read and agreed on the version of the manuscript submitted for publication.

## Ethics Statement

This study is a systematic review of published research and did not involve collecting data from human or animal participants. All included studies were conducted in accordance with their respective institutional ethical standards and the 1964 Declaration of Helsinki and its subsequent amendments. Therefore, ethical approval and informed consent were not necessary for the research.

## Supplementary Materials

**Table S1.**
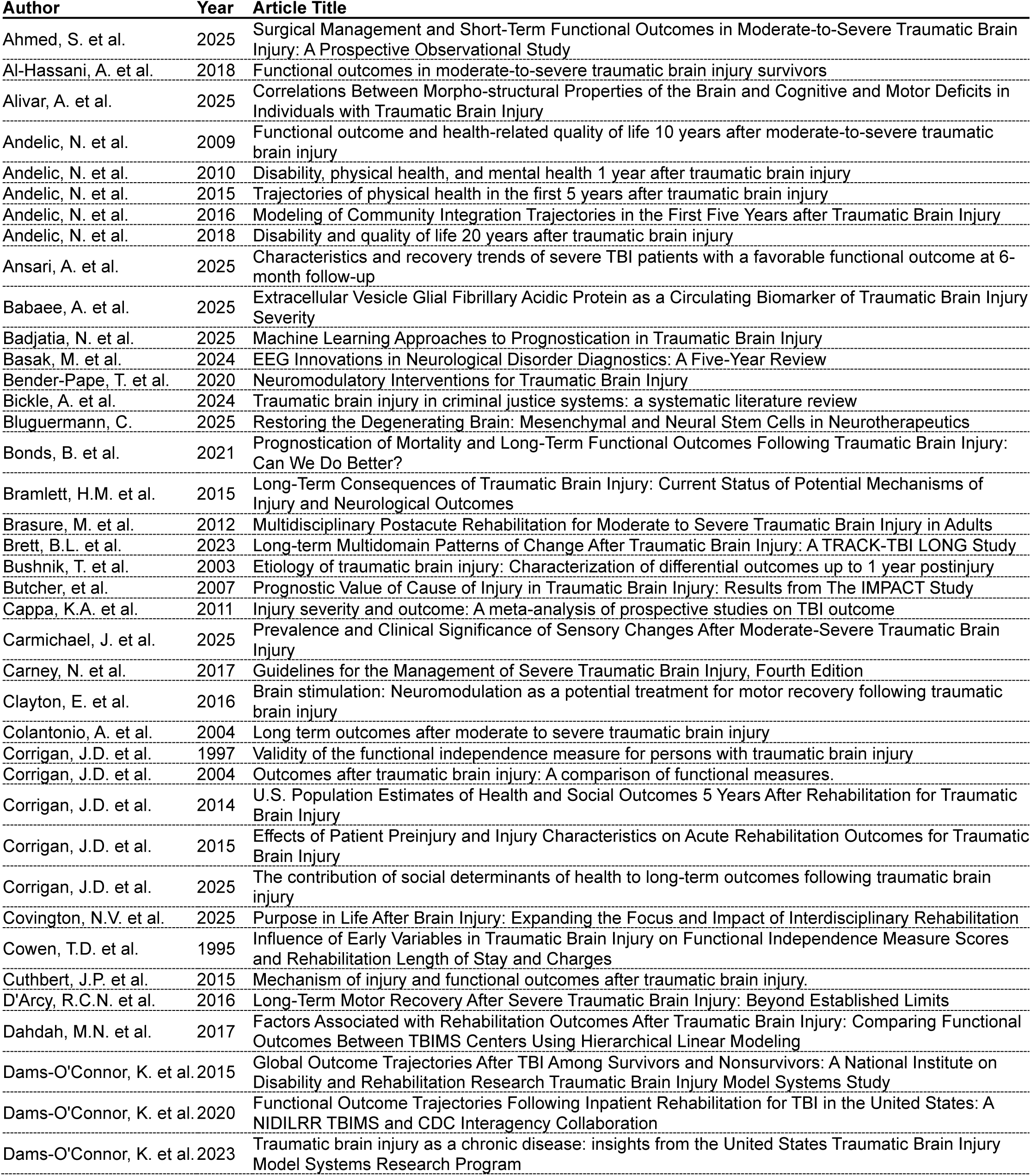
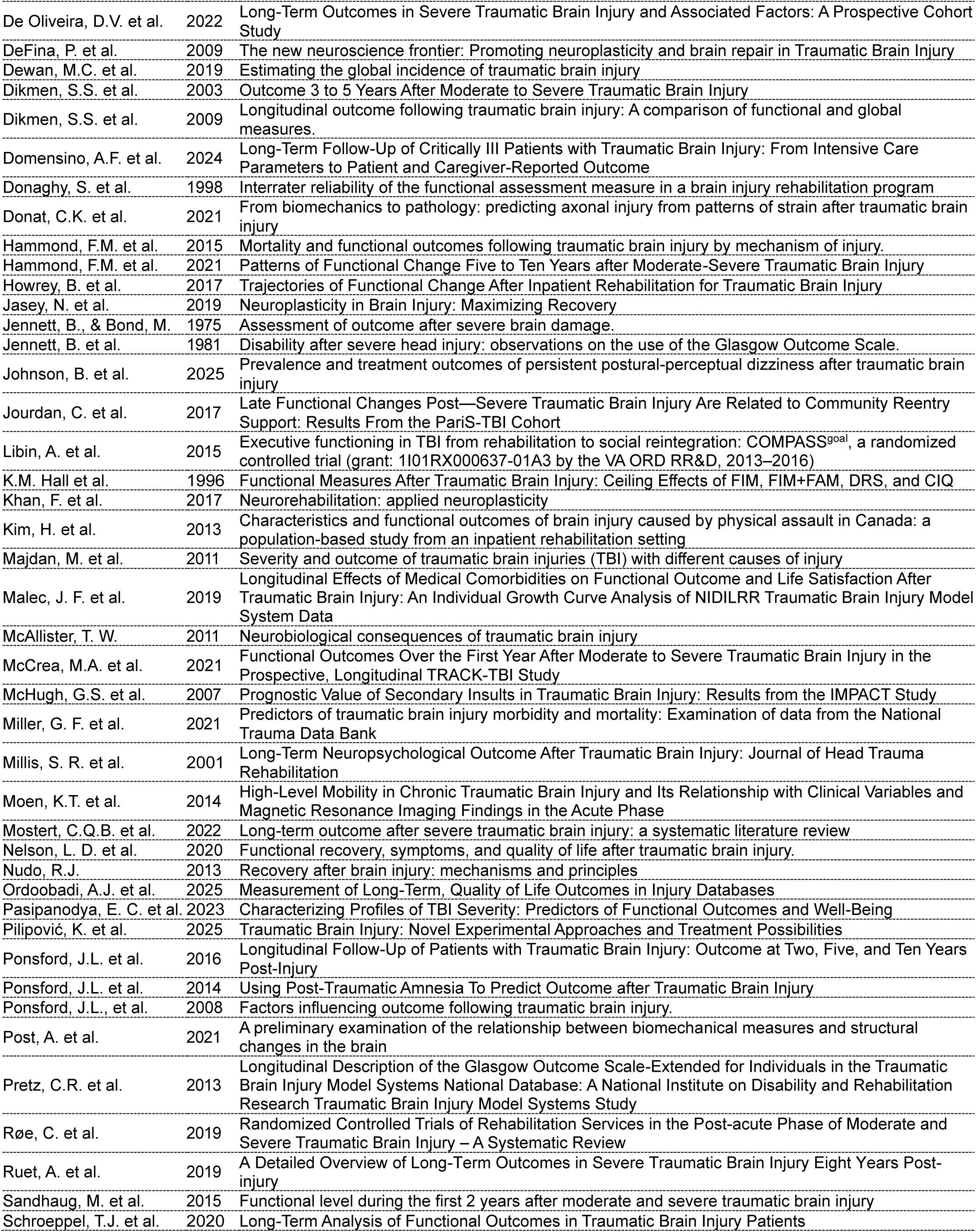
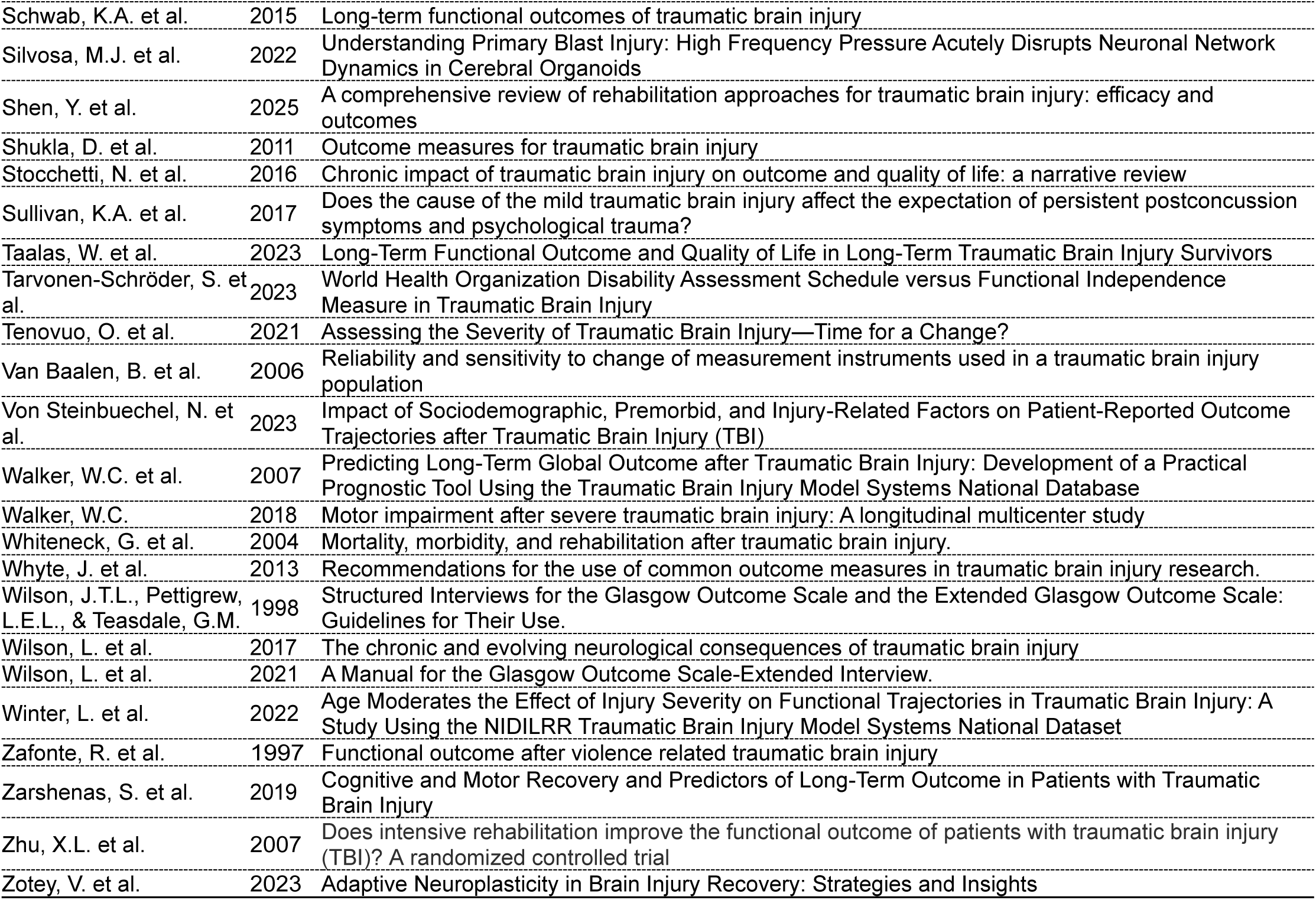
Studies Included in the Systematic Review (n = 105). Note. This table provides an overview of the 105 studies included in the systematic review, listed by first author, publication year, and article title. Collectively, the studies investigate functional outcomes, post-Traumatic Brain Injury (TBI) recovery patterns, and associated clinical, demographic, and injury-related variables across study designs and populations.

**Table S2.**
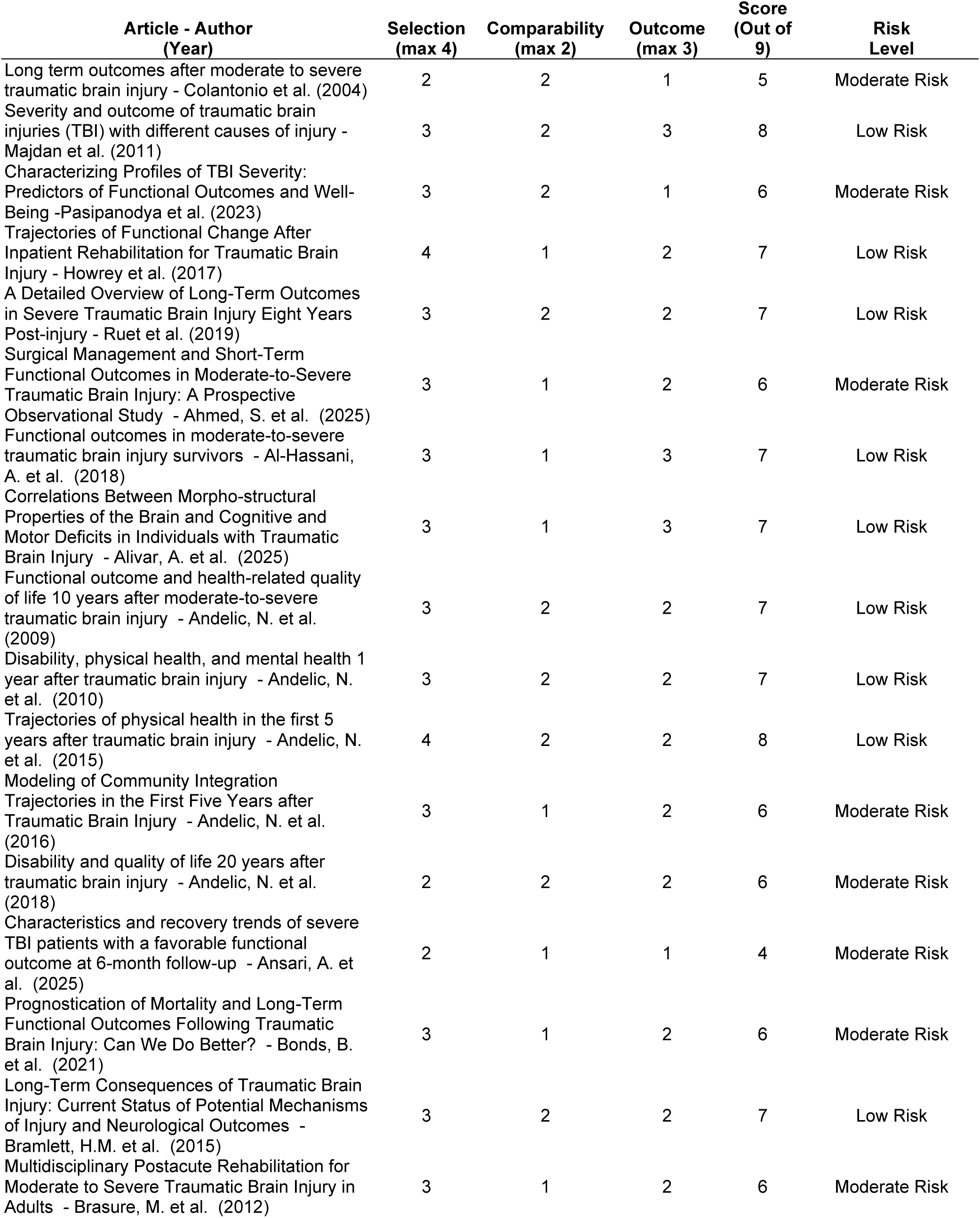
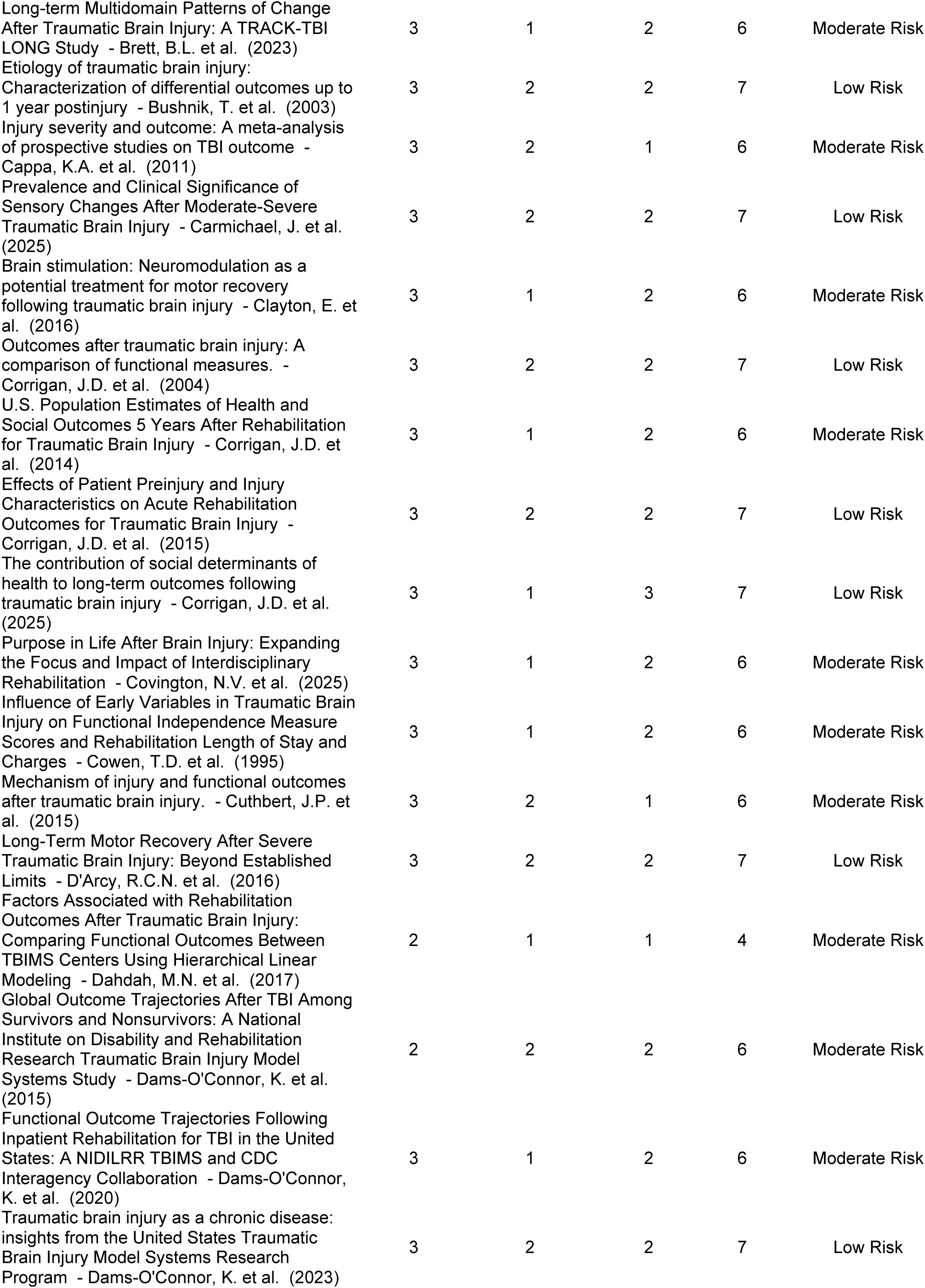
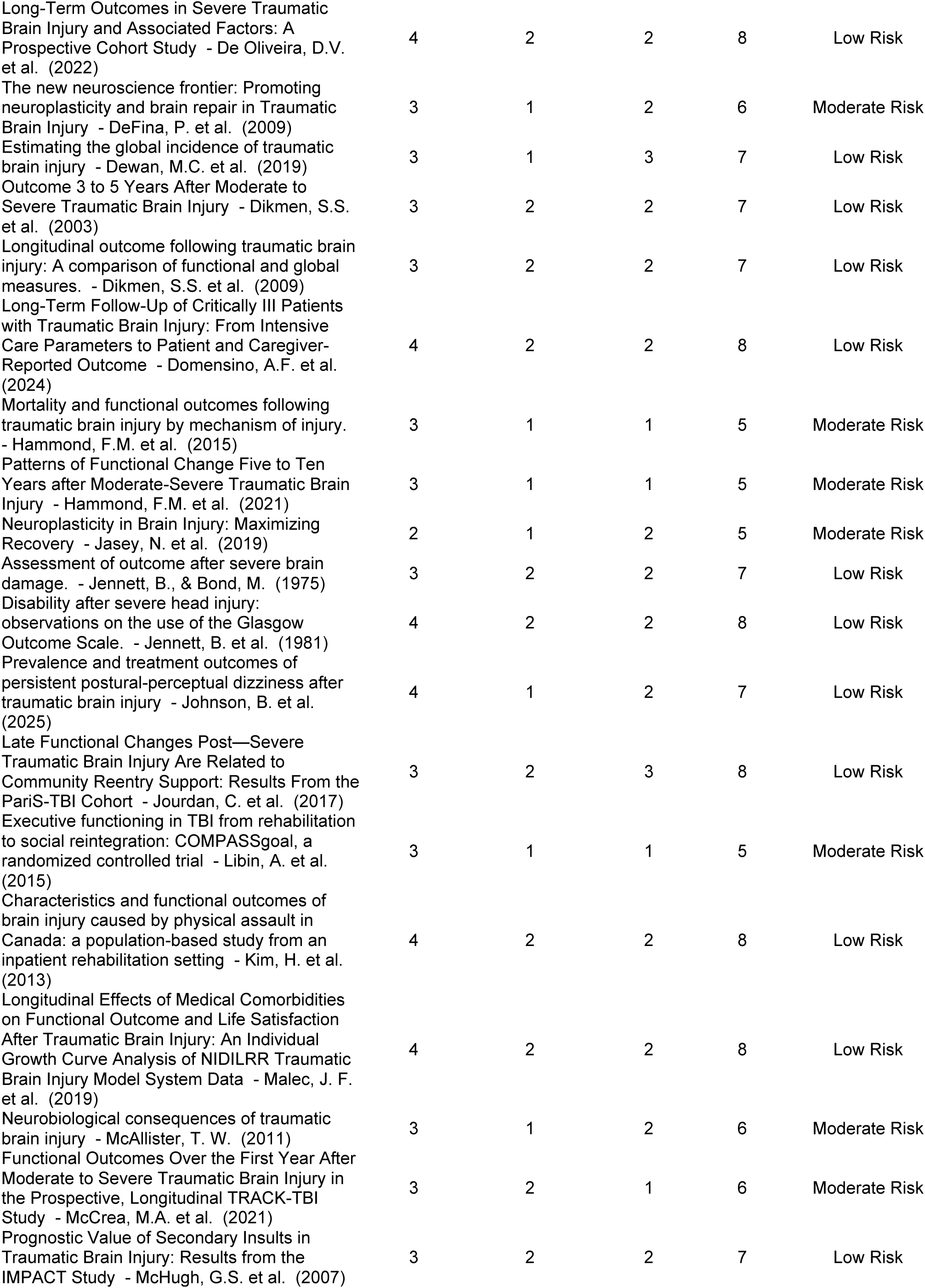
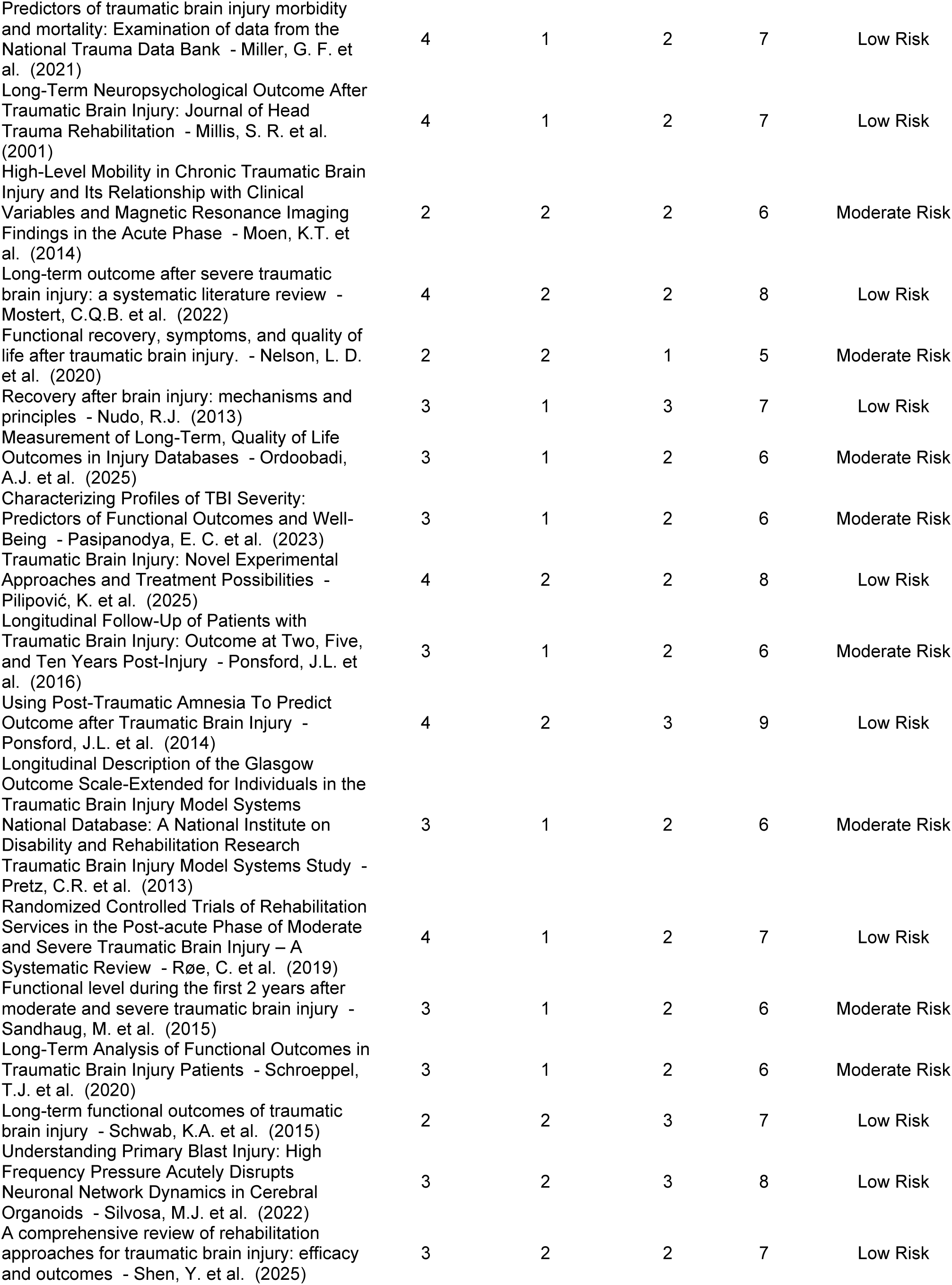
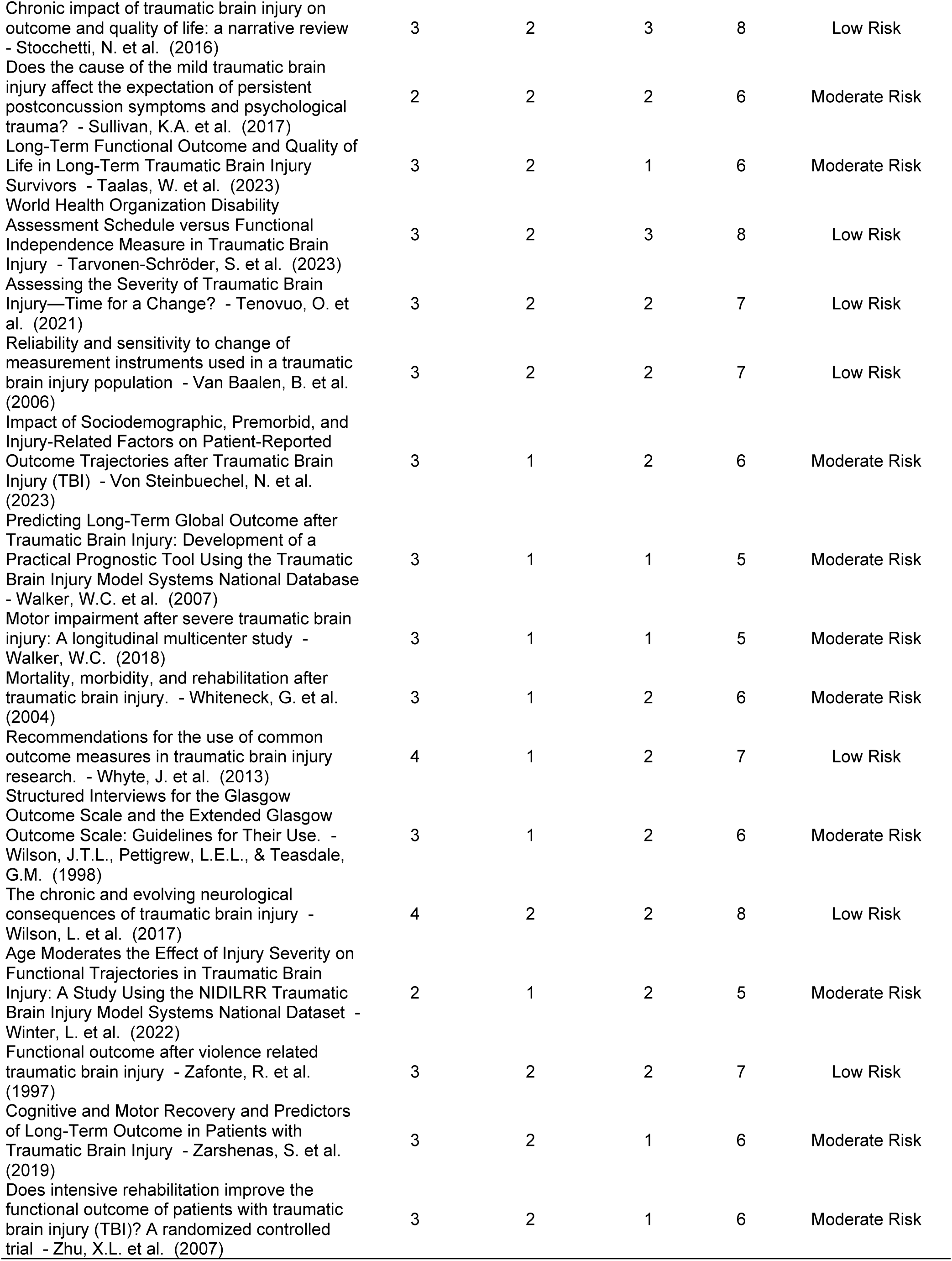
Risk of bias evaluation of observational studies using the Newcastle–Ottawa Scale (NOS) included in the systematic review.

**Table S3.** Risk of bias evaluation of randomized controlled trials (n = 2) using the Cochrane Risk of Bias Tool, version 2.0 (RoB 2) included in the systematic review.

| Article -<br>Author (Year) | Study<br>Type/Source<br>Used | Randomization | Deviations From<br>Intended<br>Interventions | Missing<br>Outcome<br>Data | Outcome<br>Measurement | Reported<br>Result | Overall<br>Rob | Rationale |
| --- | --- | --- | --- | --- | --- | --- | --- | --- |
| Executive functioning in<br>TBI from rehabilitation to<br>social reintegration:<br>COMPASS<sup>goal</sup>, a<br>randomized controlled trial<br>- Libin et al., (2015) | Study<br>protocol/trial<br>description | Some<br>concerns | Some<br>concerns | Some<br>concerns | Low RoB | Some<br>concerns | Some<br>concerns | Randomization is described, and the<br>document reports that the assessors<br>were blinded and the participants did<br>not to reveal group assignment, but<br>sequence concealment and final<br>outcome reporting were not fully<br>described in the accessible materials. |
| Does intensive<br>rehabilitation improve the<br>functional outcome of<br>patients with traumatic<br>brain injury (TBI)? A<br>randomized controlled trial<br>- Zhu et al., (2007) | Interim<br>randomized<br>controlled<br>trial report | Some<br>concerns | Some<br>concerns | Low RoB | Low RoB | Some<br>concerns | Some<br>concerns | The trial assessors were blinded, all<br>patients completed rehabilitation<br>without attrition, but allocation<br>concealment and reporting of interim<br>results were not entirely detailed in the<br>source material. |

**Table S4.** Data extraction procedures used in the systematic review. Note. The table presents detailed notes from seven representative studies. It illustrates the standardized notetaking and data extraction procedures applied to all 105 studies included in the systematic review, noting the location, study design, sample characteristics, TBI mechanisms, outcome measures, follow-up duration, and key findings.

| Article - Author (Year) | Country | Study Design | n | TBI Mechanisms | Outcome Measure(s) | Follow-up | Key Findings |
| --- | --- | --- | --- | --- | --- | --- | --- |
| Prognostic Value of Cause of Injury in Traumatic Brain Injury: Results from the IMPACT Study - Butcher et al. (2007) | United Kingdom | Meta-analysis | 8,721 | Road traffic accidents, assaults, falls, and recreational injuries | Glasgow Outcome Scale-Extended | 6 months | • There is a strong association between the cause of injury and long-term outcomes in moderate to severe TBI.<br>• Injuries from traffic accidents showed better outcomes (OR 0.66), assaults (OR 0.66), and sporting/recreational activities (OR 0.45). |
| Long term outcomes after moderate to severe traumatic brain injury – Colantonio et al. (2004) | United States | Retrospective cohort study | 306 | N/A. Different levels of injury severity | The Trail-Making Test items from the Rivermead Behavioral Memory Test, MOS SF-36 assessments of ADLs and Instrumental ADLs, and the Community Integration Questionnaire | 24 years | • Individuals were most significantly limited in activities such as managing money and shopping.<br>• Less than 30% of TBI survivors had full-time employment at the time of the follow-up, 24 years after the injury.<br>• Self-rated health correlated with many integral and fundamental ADLs. |
| Severity and outcome of traumatic brain injuries (TBI) with different causes of injury – Majdan et al. (2011) | Austria, Bosnia, Croatia, Macedonia, Slovakia | Epidemiological study | 1,109 | "Traffic-related", "falls", or "other causes." | Indicators related to survival and long-term outcomes (*specific measures were not detailed*) | 10 days post-injury to ICU discharge | • Traffic-related TBIs had the highest proportion of good recovery and only moderate disability.<br>• 52% of traffic-related TBIs had a favorable long-term outcome compared to 42% for TBIs from falls and 48% for other causes. |
| Injury Severity and Outcome: A Meta-Analysis of Prospective Studies on TBI Outcome – Cappa et al. (2011) | United States | Meta-analysis | 21,050 | N/A. Different levels of injury severity | Various measures of productivity, global disability, quality of life, independence, and global outcome. | 1 year | • Injury severity significantly predicts outcome at 1 year post-injury (average effect size $r=.257$).<br>• Higher injury severity is associated with poorer outcomes. |
| Characterizing Profiles of TBI Severity: Predictors of Functional Outcomes and Well-Being – Pasipanodya et al. (2023) | United States | Longitudinal study | 379 | Violence, sports, motor vehicles, falls, and others | Length of stay, Functional Independence Measure, Disability Rating Scale, Glasgow Outcome Scale-Extended, and the Satisfaction with Life Scale | Hospital discharge and 1-year post-injury | • There are significant differences in length of stay, FIM, and DRS scores between the LPA classes, with class 4 experiencing longer stays and lower FIM and DRS scores.<br>• LPA class 4 exhibited worse functional recovery and well-being outcomes than other classes.<br>• Members of LPA class 4 sustained the most severe injuries. |
| Trajectories of Functional Change After Inpatient Rehabilitation for Traumatic Brain Injury - Howrey et al. (2017) | United States | Prospective study | 16,583 | N/A. Different levels of injury severity | Functional Independence Measure | Hospital discharge and follow-ups (103 days on average) | • Open head injuries and comorbidities are associated with a poorer motor recovery trajectory. |
| A Detailed Overview of Long-Term Outcomes in Severe Traumatic Brain Injury Eight Years Post-injury – Ruet et al. (2019) | France | Prospective study | 504 | Road traffic accidents and falls | Hospital Anxiety and Depression Scale, Glasgow Outcome Scale-Extended, Brain Injury Complaint Questionnaire, Dysexecutive Questionnaire. | 8 years | • 37% of participants had moderate disability after 8 years.<br>• 10.5% of participants had anxiety disorder, 9.2% of participants had depression, and 14.5% of participants had both.<br>• Longer education, a shorter ICU stay, higher GCS scores at discharge, and higher GOSE scores at 1-year post-injury were associated with improved outcomes at 8 years. |

**Table S5.** Summary of Standardized Functional Outcomes (zFIM and zGOSE) of individuals stratified by their respective TBI Mechanisms, including means, standard deviations, ranges, and 95% confidence intervals. *MVAs = Motor Vehicle Accidents*

| TBI Mechanism | k | Z<sub>FIM</sub><br>Range | Z<sub>GOSE</sub><br>Range | Z<sub>FIM</sub><br>Mean | Z<sub>FIM</sub><br>SD | Z<sub>FIM</sub><br>95% CI<br>(L) | Z<sub>FIM</sub><br>95% CI<br>(U) | Z<sub>GOSE</sub><br>Mean | Z<sub>GOSE</sub><br>SD | Z<sub>GOSE</sub><br>95% CI<br>(L) | Z<sub>GOSE</sub><br>95% CI<br>(U) | Pooled<br>Mean z | Pooled<br>95% CI<br>(L) | Pooled<br>95% CI<br>(U) |
| --- | --- | --- | --- | --- | --- | --- | --- | --- | --- | --- | --- | --- | --- | --- |
| Falls | 82 | 0.87–1.28 | 0.00–0.63 | 1.08 | 0.10 | 1.06 | 1.10 | 0.32 | 0.16 | 0.29 | 0.35 | 0.70 | 0.66 | 0.74 |
| MVAs | 79 | 1.64–2.08 | 0.63–1.88 | 1.86 | 0.11 | 1.84 | 1.88 | 1.26 | 0.31 | 1.19 | 1.33 | 1.56 | 1.52 | 1.60 |
| Penetrating | 92 | –2.07––0.23 | –1.88––0.42 | –1.15 | 0.46 | –1.24 | –1.06 | –1.15 | 0.37 | –1.23 | –1.07 | –1.15 | –1.21 | –1.09 |
| Assaults | 88 | –1.11–0.62 | –0.63–0.63 | –0.25 | 0.43 | –0.34 | –0.16 | 0.00 | 0.32 | –0.07 | 0.07 | –0.12 | –0.18 | –0.06 |
| Recreational | 81 | 1.81–2.13 | 1.25–1.88 | 1.97 | 0.08 | 1.95 | 1.99 | 1.57 | 0.16 | 1.54 | 1.60 | 1.77 | 1.74 | 1.80 |

**Figure S1.**
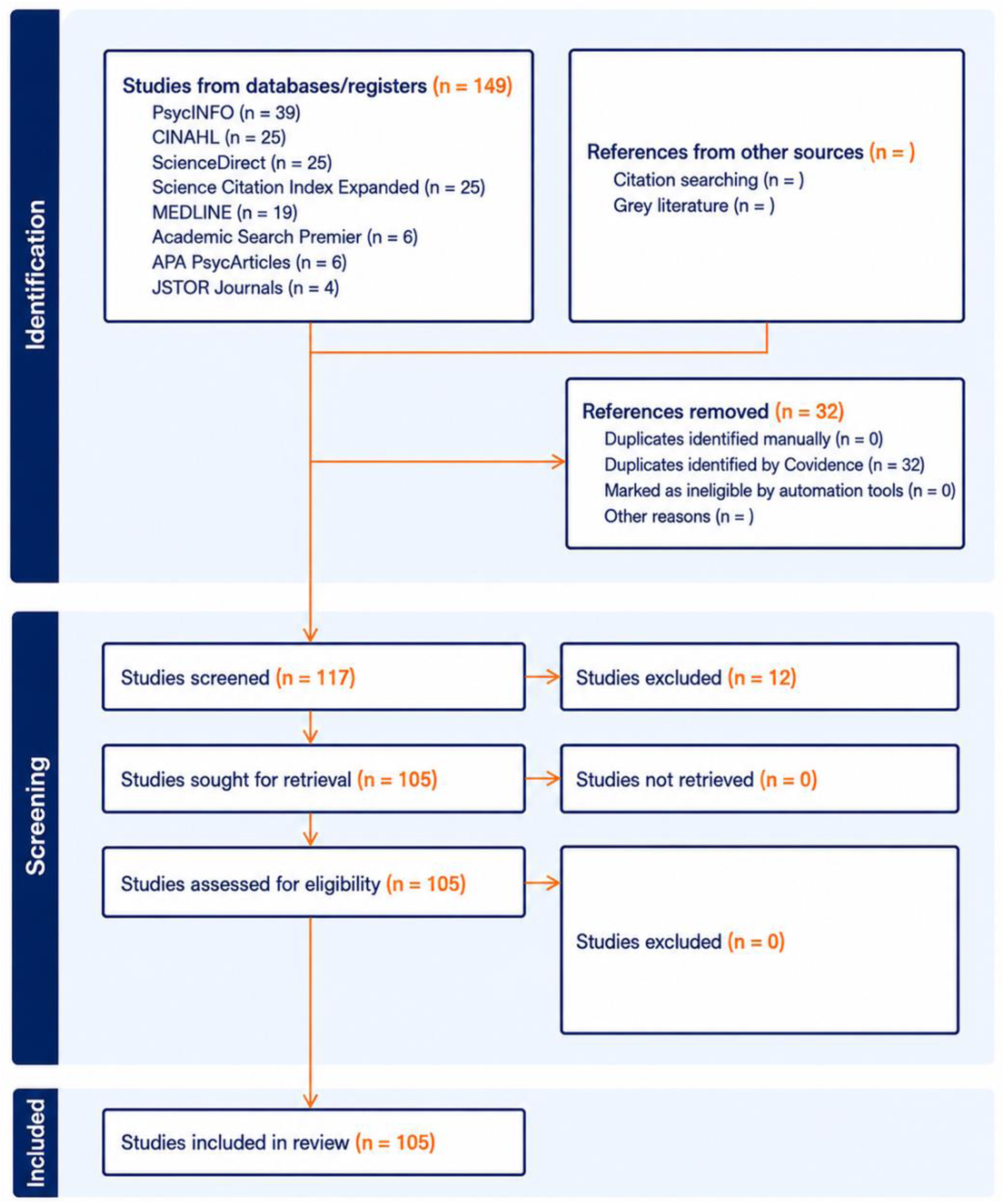
Preferred Reporting Items for Systematic Reviews and Meta-Analyses (PRISMA) flow diagram depicting the study selection process. The diagram illustrates the four phases of the review—identification, screening, eligibility evaluation, and inclusion—specifying the number of records identified (n = 149), screened (n = 117), excluded (n = 12), and finally included (n =105) in the systematic review.

**Figure S2.**
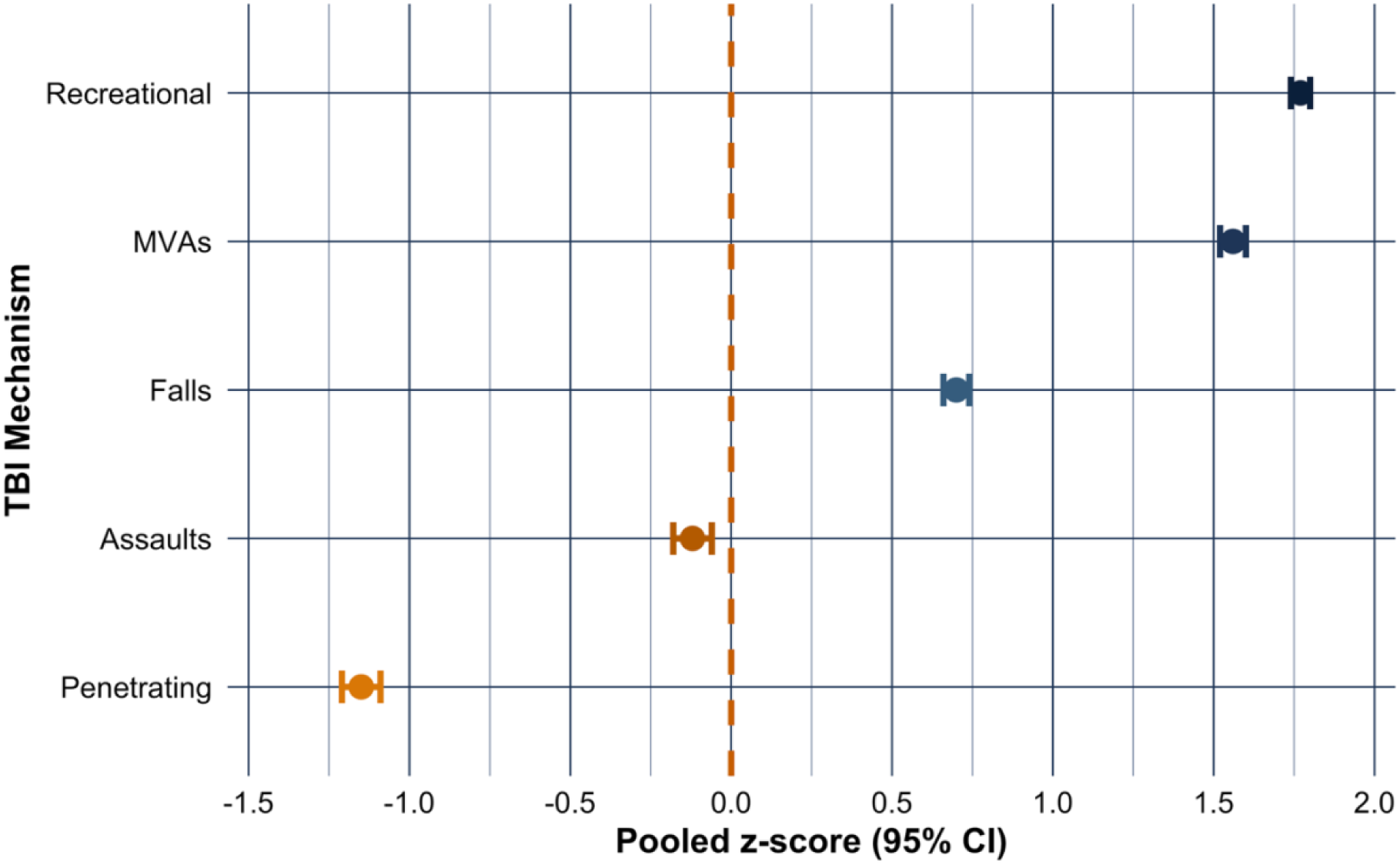
Forest plot of pooled standardized functional outcomes (z_FIM_ and z_GOSE_) of individuals stratified by their respective mechanisms of Traumatic Brain Injury (TBI). The points indicate pooled z-score estimates for each TBI mechanism, and error bars represent 95% confidence intervals. The dashed vertical line at zero serves as a reference value, indicating that values to the right of the line represent superior functional outcomes and those to the left represent poorer functional outcomes. *\*MVAs = Motor Vehicle Accidents\**

